# Experience of Saudi Dental Practitioners with Intraoral Scanners

**DOI:** 10.1101/2022.07.10.22277476

**Authors:** Ibrahim K. Al-Ibrahim, Anfal H. Alotaibi, Alanoud S. Alshammari, Ahmed A. Madfa

**Affiliations:** Department of Restorative Dental Science, Collage of Dentistry, University of Ha’il, Ha’il, Kingdom of Saudi Arabia; Dental Intern, Collage of Dentistry, University of Ha’il, Ha’il, Kingdom of Saudi Arabia

**Keywords:** Digital Application, Digital impression, Intraoral scanner, Saudi Arabia

## Abstract

**Background:** The objective of this study was to investigate the experience of Saudi dental practitioners with intraoral scanners, investigate the current knowledge and improve the practice accordingly.

**Methods:** At random, electronic surveys were distributed to Saudi dental practitioners. The study received 400 questionnaires, with 310 judged valid for the study. The questionnaire was divided into the following sections: (i) Practitioners’ demographic information such as gender, practice level, and practice experience. (ii) Experience and benefits of intraoral scanners. (iii) Require skills and training of IOS. (iv) Knowledge of IOS usage. Descriptive statistics such as numbers and percentages were used to analyze the collected data. The Chi-square and Fisher’s Exact tests were used to assess the results.

**Results:** There were 161 women (51.8%) and 149 men among the participants (47.9%). General practitioners (198, or 63.7%) had the most subjects, followed by specialists (80, or 25.7%) and consultants (32, or 10.3%). In terms of IOS use in dental practice, most participants (70.6%) do not use it, while less than one-third do. The majority of participants (52.3%) intend to purchase IOS with significant variations based on gender, experience, and level of practice (p<0.05). Compared to traditional, most participants believe that IOS will eventually replace it, improve quality, and be more aesthetically pleasing. Most dentists believe that using IOS requires special skills and training. More than half of dentists believe IOSs have the same level of accuracy as conventional in producing three units FPDs, implant prosthesis, and complete denture.

**Conclusion:** It can be concluded that dentists have a high level of satisfaction and a favorable attitude toward using IOS technology in clinical dentistry practice.

## Background

Since the introduction of computer-aided design and computer-aided manufacturing (CAD/CAM), digital technology in dentistry has advanced rapidly.^(1)^ Digital impressions are taken in the CAD/CAM system using an intraoral scanner.^(2)^ Intraoral scanners (IOSs) are used to digitalize teeth, an effective method for obtaining a direct digital model from the patient.^(3)^ IOS captures information about projecting light in the same way as the camera does.^(2)^ Therefore, the light reflection times of the subject surface are measured by the intraoral scanner.^(2)^ For scanning, intraoral cameras use either video or still photography. Furthermore, a 3D image can be created by combining several still images.^(2)^ These are the fundamentals, and each manufacturer employs its techniques on top of them, and intraoral cameras can employ various methods for data collection.^(2)^

Digital impressions and scanning technologies provide several advantages, including improved patient acceptability, less impression material distortion, and possible cost and time savings.^(4)^ Furthermore, digital impressions offer easy impression repeatability, direct model visualization, and chairside production for CAD/CAM restorations.^(3)^ Additionally, IOS decreases patient anxiety and discomfort. Many patients today suffer from anxiety and have a strong gag reflex, making traditional impressions impossible to tolerate; in these cases, using light to replace trays and materials is an excellent alternative.^(5)^ Furthermore, IOS save time for the dentist and can simplify clinical procedures, particularly for complex impressions.^(5)^ Moreover, it provides the ability to save and transfer digital images between the dental office and the laboratory indefinitely.^(6)^

A learning curve is a visual representation of the learning rate over time or in multiple instances.^(7)^ Many studies in general medicine have looked into how new technologies affect system users’ learning curves.^(8-12)^ However, there have been few studies on the learning curve in dentistry.^(13-16)^ Furthermore, there have not been any systematic or randomized clinical trials evaluating the digital impression learning curve.^(17)^ When considering the purchase of a new system, a practising dentist must understand the learning curve of digital impressions and the scanner’s applicability.^(17)^ According to Birnbaum and Aaronson^(18)^, users of digital intraoral scanners would need time and education to learn new skills to provide a quick and reliable restoration with the best fit. The learning process is reflected in the decrease in the time required to take digital impressions and the number of virtual model images.^(17)^ Enhancing dentists ability to use a certain technology or therapeutic procedure can help save time in the chair while also improving the quality of the treatment.^(19)^ Additionally, the trueness of the scanned images of the single-image-based system was affected by repeated experience, clinical experience, and the scanned region.^(19)^ As a result, users require multiple practice opportunities for effective clinical application.^(19)^ Although dentists may not afford to introduce every new technology, newer versions appear to be more accurate and easier to apply in clinical practice.^(19)^ Resende et al. concluded that operator experience, IOS type, and scan size all influenced the accuracy of intraoral scans.^(1)^ Therefore, scans were more accurate due to more experienced operators and smaller scan sizes.^(1)^ Additionally, more experienced dentists performed faster scans.^(1)^

Dental models must be highly accurate to ensure a proper fit of dental restorations and correct virtual articulation.^(3)^ The accuracy of any dental impression method is determined by trueness and precision.^(2)^ The deviation of the tested impression method from the original geometry determines trueness, whereas differences within the same test group determine precision.^(2)^ Ahlholm et al., in a systematic review^(2)^, concluded that, in the fabrication of crowns and short fixed partial dentures (FPDs), digital impression accuracy is comparable to that of conventional impression methods. Thus, both techniques can be used. Furthermore, digital imprint systems produce a clinically acceptable fit for implant-supported crowns and FDPs; however, for big, full-arch FPDs, the traditional impression approach produces superior accuracy than the digital method.^(2)^ Therefore, the traditional method may be preferred for full arch impression.

Intraoral scanners usage has been a focus of attention of many researchers due to its importance in future practice. Unfortunately, there is no similar study conducted in Saudi Arabia. For this reason, it is crucial to conduct a study in intraoral scanners usage among dental practitioners in Saudi Arabia to investigate the current situation and fill the knowledge gap in the critical area and give starts for further investigations. Furthermore, that will help academicians determine the existing knowledge and improve the future outcomes. Therefore, this study aimed to investigate the experience of Saudi dental practitioners with intraoral scanners, investigate the current knowledge and improve the practice accordingly.

## Method

A cross-sectional study of Saudi dental practitioners was conducted. The University of Hail’s Ethical Committee approved the study, and it was carried out following the Helsinki Declaration’s principles. The confidentiality of the subject was strictly followed. According to the OpenEpi® sample size calculator, the study’s estimated sample size was 300, with a power of 84 percent and a P=5%. This study includes dental practitioners.

The study sample consists of 600 Saudi Arabian practitioners selected at random and given a self-explanatory questionnaire. Practitioners were given questionnaires to complete based on their observations and experiences. Before the data was collected, participants gave their informed consent.

There were 400 questionnaires returned, with 310 judged valid for the study. The survey and a cover letter emphasizing that all responses will be treated anonymously were emailed to all selected dental practitioners. Non-responders were automatically sent four reminders at one-week intervals using the survey technology. The surveys were given out from July to September 2021.

A self-administered structured questionnaire with 13 questions was used to perform a pilot survey with a convenient sample of 20 dental practitioners. Based on their feedback, no adjustments to the questionnaire were required. Electronic surveys were delivered to Saudi dental practitioners at random. The questionnaire was divided into the following sections:

1. Practitioners’ demographic information such as gender, practice level, and practice experience.
2. Experience and benefits of intraoral scanners.
3. Require skills and training of IOS.
4. Knowledge of IOS usage.

Frequencies, Crosstabs, Chi-Square, and Fisher’s Exact tests were used to determine the significance of gender, practice level and experience differences in the knowledge of intraoral scanners among Saudi dental practitioners. The completed questionnaires were entered in Windows Excel and statistically analyzed with the Social Sciences software version 28 (IBM SPS Statistics). The acquired data were analyzed using descriptive statistics such as numbers and percentages.

## Results

A total of 311 dentists completed the questionnaire. There were 161 women (51.8%) and 149 men among the participants (47.9%). General practitioners (198, or 63.7%) had the most subjects, followed by specialists (80, or 25.7%) and consultants (32, or 10.3%).

The distribution of questions and scores related to IOS knowledge is shown in Table 1. Gender, experience, and practice level impact IOS knowledge, as shown in Tables 2, 3, and 4. IOS was unknown to 180 (58.1%) of 311 dentists, while it was known to 130 (41.9%). Men and women have no difference in perceptions of IOS (p >0.05), as shown in Table 2. Consultants and specialists had significantly higher levels of knowledge than general practitioners (p <0.01), as shown in Table 4. Moreover, as shown in Table 3, dentists with more than ten years of experience and dentists with five to ten years of experience better understood IOS than dentists with less than five years of experience (p<0.01).

In terms of IOS use in dental practice, most participants (70.6%) do not use it, while less than one-third do. Tables 3 and 4 demonstrate that the usage of IOS in dental offices varies significantly based on experience and level of practice (p>0.01), whereas gender shows no difference (p>0.05). Furthermore, the majority of participants (52.3%) intend to purchase IOS with significant variations based on gender, experience, and level of practice (p<0.05). Figure1 shows how intraoral scanners improves clinical practice.

**Figure 1.**
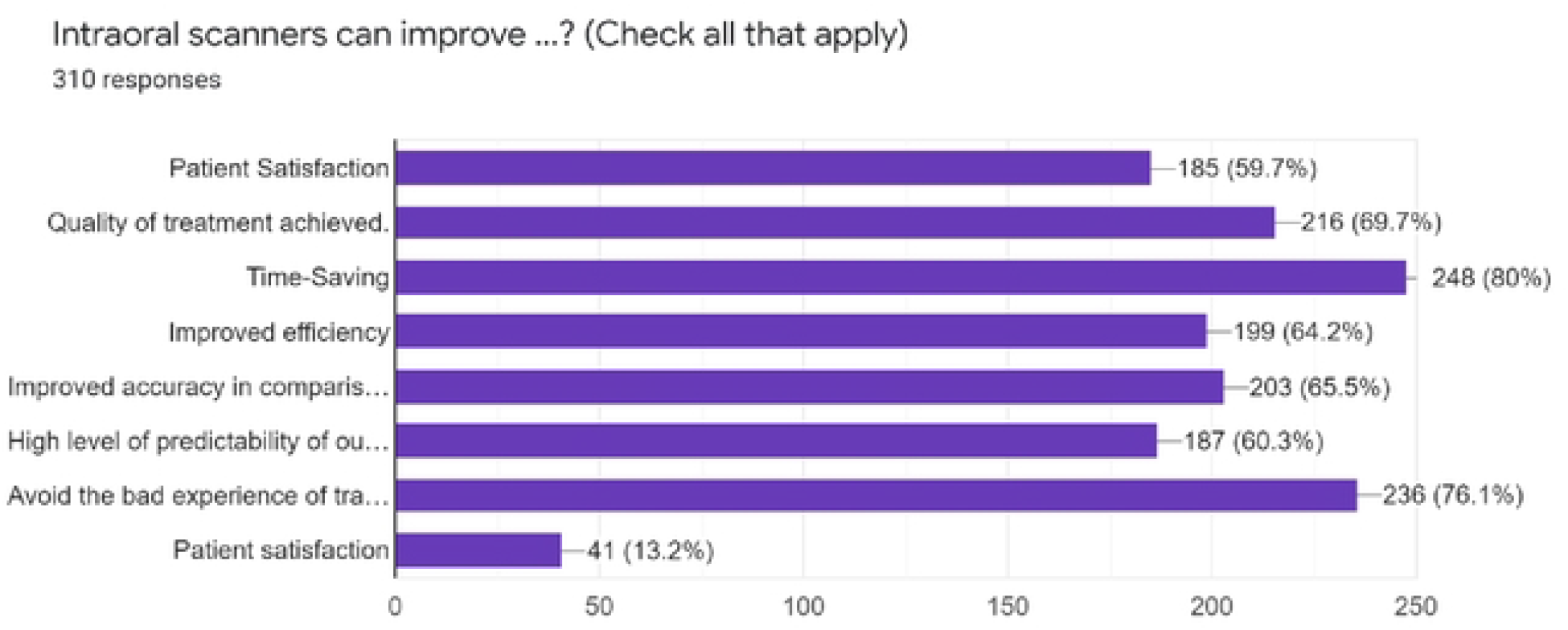
shows how intraoral scanners improves clinical practice.

Compared to traditional, most participants believe that IOS will eventually replace it, improve quality, and be more aesthetically pleasing. According to the gender, experience, and practice level of dentists, there were no statistically significant relationships between the perception of IOS to replace traditional procedures, improve quality, and enhance aesthetics (p>0.05).

Most dentists believe that using IOS requires special skills and training. In terms of training, there were no significant differences in gender or practice level of dentists (p>0.05); however, experience shows considerable differences(p<0.01), as shown in Tables 2,3 and 4.

More than half of dentists believe IOSs have the same level of accuracy as conventional in producing three units FPDs, implant prosthesis, and complete denture. The experience of dentists was statistically significant in the use of IOSs for the manufacturing of three units FPDs (p<0.05), whereas gender and practice level were not (p>0.05). In terms of using IOSs for implant prosthesis, experience, and practice level show significant differences (p<0.05), while gender did not (p>0.05). Furthermore, the level of training varies significantly in using IOSs to produce complete dentures; however, gender and experience of dentists show no significant relationship (p>0.05).

## Discussion

In today’s clinical practice, digital dental technology is unavoidable. Dental students are now growing up in a digital world, which influences their preferences, expectations, and acquiring new knowledge. Dentists are increasingly interested in incorporating digital technology into teaching and learning. Furthermore, the rapid development of digital technology poses a challenge for teachers, necessitating constant adjustments and changes in the curriculum. Dentists are confronted with a plethora of digital technologies. The focus of this study was on their acceptance and rejection, emphasising the barriers and motivating factors encountered along the way.^(20)^ These extend far beyond the technical capabilities of digital dental technologies. Adoption of dental technologies varies depending on the individual and the technology, but overall trends can be identified, which should be investigated further in future research. Many clinics worldwide are using intraoral scanners because their use is rapidly expanding. As a result, the purpose of this study was to determine dentists’ knowledge and attitudes toward intraoral scanners in Saudi Arabia.

The grown interest in intraoral scanners has increased in research volume. Many researchers have chosen to focus on the accuracy and time efficiency of intraoral scanners, as well as the perspectives of patients, dentists, students, and assistants.^(21-23)^

In the present study, more than a quarter of the dentists (29.4%) indicated using intraoral scanners in their clinics. Essentially, user acceptance of intraoral scanners is determined by scanner user experience.^(24, 25)^ Many dentists refuse to use these new tools due to the lengthy learning curve. They believe that learning intraoral scanning will be just as difficult for a student or newly graduated dentist as traditional impression taking.^(18)^ Investigating how intraoral scanners are used is a critical step in integrating them into daily clinical practice.

On the level of satisfaction and attitude, the current survey findings reflect overall satisfaction and a positive attitude among participating dentists toward the use and outcome of intraoral scanners in clinical practice. Efficiency was defined by Lee et al. as time and number of retakes/rescans.^(21)^ Among other clinical benefits, IOS is more efficient than impression making because it eliminates the need for time spent mixing and setting impression material, disinfecting the impression, and pouring stone models.^(26, 27)^ Areas that are less than ideal can easily be removed to allow for recapture or simply re-scanning.^(28)^

On the other hand, critical errors in the impressions (e.g., bubbles along preparation margins, unclear margins) will render the entire impression useless, and the whole impression will need to be made again.^(29)^ The IOS and impression making are likely to necessitate distinct skill sets.^(17, 21, 23)^ Dentists in this study were exposed to various intraoral scanners; differences in the system and scanning protocols may result in variations in impression making success.^(30, 31)^

There is a learning curve for implementing IOS in the dental clinic, and this must be taken into account.^(29)^ Subjects who have a strong interest in technology and computers (for example, young dentists) will find it very easy to incorporate IOS into their practice. In the current study, most of the surveyed dentists stated that using IOS requires skills and training. Intraoral scanners, like any other impression technique, have a learning curve.^(17)^ Some mouth areas, such as the distal surfaces of the last tooth in an arch and the proximal surfaces near a bounded saddle, may be challenging to capture with intraoral scanners.

For beginners, moving the tip of intraoral scanners around while following the scanner’s signals based on previously captured surfaces is difficult.^(30)^ The IOS and impression making are likely to necessitate distinct skill sets.^(23, 32)^ Dentists in this study were exposed to various intraoral scanners; differences in the system and scanning protocols may result in variations in impression making success.^(31)^

The clinician and dental technician can assess the quality of the impression in real-time using IOS.^(33)^ The dentist can e-mail the scan to the laboratory immediately after it is completed, and the technician can accurately check it.^(33, 34)^ Suppose the dental technician is not satisfied with the quality of the received optical impression. In that case, they can immediately request that the clinician make another one without wasting time or calling the patient for a second appointment. ^(33, 34)^ This feature facilitates and strengthens communication between the dentist and dental technician. ^(33, 34)^

According to the results of this survey, a significant number of responding dentists believed that IOS could replace the traditional method. In the case of single tooth restoration and fixed partial prostheses of up to 4–5 elements, the scientific literature considers optical impressions to be clinically satisfactory and comparable to conventional impressions.^(23)^ In fact, for these types of short-span restorations, the trueness and precision obtained with optical impressions are comparable to those obtained with conventional impressions.^(35, 36)^ However, in the case of long-span restorations such as partial fixed prostheses with more than five elements or full-arch prostheses on natural teeth or implants, optical impressions do not appear to be as accurate as conventional impressions. ^(35, 36)^ The error generated during intraoral scanning of the entire dental arch does not appear to be compatible with the fabrication of long-span restorations, which still require conventional impressions. ^(35, 36)^

The current study has limitations because it is only cross-sectional, and clinical studies are undoubtedly required to draw more specific conclusions about the accuracy and clinical indications of IOS in prosthetic and implant dentistry and orthodontics. More randomised controlled trials on IOS are needed to perform a systematic review of the literature based on an adequate number of cases/patients treated effectively. A clinical scenario could provide additional information considering the limitations and complications of impression making at the start of their clinical practice, so the lack of an actual patient practice should be considered.

The findings of this study can be extrapolated to dentists with more clinical experience and who should be more efficient in impression making; thus, a higher proportion of them are expected to prefer impression making over IOS. More research is needed to investigate dentists’ preferences and perceptions of IOS and impression making.

## Conclusions

Despite the limitations of the current study, it can be concluded that dentists have a high level of satisfaction and a favorable attitude toward the use of IOS technology in clinical dentistry practice. In Saudi Arabia, dentists prefer IOS. While intraoral scanning appears to have advantages, many students still prefer traditional impression making because it is more efficient. Dental professionals should offer more IOS courses as part of their continuing education modules. Meanwhile, dental schools should prepare students to be proficient in both techniques to deal with various clinical situations.

In routine dental care, digital tools and applications are now widely used. As a result, future dentists must be prepared for their daily work lives by considering the trend toward digitization and ongoing developments in dental curricula. There is a need to develop universally accepted digital education standards, at least among dental universities within individual countries.

## Data Availability

All relevant data are within the manuscript and its Supporting Information files

## Declarations

### Ethics approval and consent participate

The Ethics Committee of Scientific Research, University of Hail, Saudi Arabia, approved the protocol of this study. All methods were performed in accordance with the declaration of Helsinki. (Ethical number: H-2021-148). Participants obtained informed consent before the data was collected.

### Consent for publication

“Not Applicable”.

### Availability of Data and Materials

The data that support the results of this study are accessible from the corresponding author upon reasonable request.

### Authors’ contributions

IKA and AAM contributed to the concept of the research, study design, statistical analysis, writing the original draft, and reading and editing the final paper. AHA and ASA contributed to data gathering and statistical analysis. The final manuscript was reviewed and approved by all writers.

## Acknowledgements

Not applicable.

